# Long-term development of white matter fibre density and morphology up to 13 years after preterm birth

**DOI:** 10.1101/2020.04.01.20049585

**Authors:** Claire E Kelly, Deanne K Thompson, Sila Genc, Jian Chen, Joseph YM Yang, Chris Adamson, Richard Beare, Marc L Seal, Lex W Doyle, Jeanie LY Cheong, Peter J Anderson

**Affiliations:** Victorian Infant Brain Study (VIBeS), Murdoch Children’s Research Institute, Melbourne, Australia; Developmental Imaging, Murdoch Children’s Research Institute, Melbourne, Australia; Department of Paediatrics, The University of Melbourne, Melbourne, Australia; Florey Institute of Neuroscience and Mental Health, Melbourne, Australia; Cardiff University Brain Research Imaging Centre (CUBRIC), Cardiff University, Cardiff, UK; Department of Neurosurgery, The Royal Children’s Hospital, Melbourne, Australia; Neuroscience Research, Murdoch Children’s Research Institute, Melbourne, Australia; Newborn Research, The Royal Women’s Hospital, Melbourne, Australia; Department of Obstetrics and Gynaecology, The University of Melbourne, Melbourne, Australia; Turner Institute for Brain and Mental Health, Monash University, Melbourne, Australia

## Abstract

**Background:** It is well documented that infants born very preterm (VP) are at risk of brain injury and altered brain development in the neonatal period, however there is a lack of long-term, longitudinal studies on the effects of VP birth on white matter development over childhood. Most previous studies were based on voxel-averaged, non-fibre-specific diffusion magnetic resonance imaging (MRI) measures, such as fractional anisotropy. In contrast, the novel diffusion MRI analysis framework, fixel-based analysis (FBA), enables whole-brain analysis of microstructural and macrostructural properties of individual fibre populations at a sub-voxel level. We applied FBA to investigate the long-term implications of VP birth and associated perinatal risk factors on fibre development in childhood and adolescence.

**Methods:** Diffusion images were acquired for a cohort of VP (born <30 weeks’ gestation) and full-term (FT, ≥37 weeks’ gestation) children at two ages: mean (SD) 7.6 (0.2) years (*n*=138 VP and 32 FT children) and 13.3 (0.4) years (*n*=130 VP and 45 FT children). 103 VP and 21 FT children had images at both ages for longitudinal analysis. At every fixel (individual fibre population within an image voxel) across the white matter, we compared FBA metrics (fibre density (FD), cross-section (FC) and a combination of these properties (FDC)) between VP and FT groups cross-sectionally at each age, and longitudinally between ages. We also examined associations between perinatal risk factors and FBA metrics in the VP group.

**Results:** Compared with FT children, VP children had lower FD, FC and FDC throughout the white matter, particularly in the corpus callosum, tapetum, inferior fronto-occipital fasciculus, fornix and cingulum at ages 7 and 13 years, as well as the motor pathways at age 13 years. VP children also had slower FDC development in the corpus callosum and corticospinal tract between ages 7 and 13 years compared with FT children. Within VP children, earlier gestational age at birth, lower birth weight z-score, and neonatal brain abnormalities were associated with lower FD, FC and FDC throughout the white matter at both ages.

**Conclusions:** VP birth and concomitant perinatal risk factors are associated with fibre tract-specific alterations to axonal development in childhood and adolescence.

## Introduction

Preterm birth is a significant clinical and public health concern given the large and increasing number of infants who survive with serious health and developmental problems (approximately 11% of births are preterm at <37 weeks’ gestation and approximately 1.8% are very preterm (VP) at <32 weeks’ gestation).^1,2^ Preterm infants are susceptible to brain injury, particularly white matter injury, and subsequent disruptions to the normal development of both the white matter and grey matter.^3^ The manifestations of brain injury and altered brain maturation can be assessed using magnetic resonance imaging (MRI), with diffusion-weighted MRI providing an unprecedented method for studying white matter on the microscopic scale. White matter microstructural disturbances in VP infants have clinical relevance due to their association with a range of motor, cognitive and behavioural delays.^4^

A major challenge in analysing diffusion images is accounting for the complex, crossing fibre geometries in the white matter, which are present in up to 90% of white matter image voxels.^5^ The most common diffusion image analysis approach, diffusion tensor imaging (DTI), cannot resolve crossing fibres, meaning the microstructural parameters derived from DTI (fractional anisotropy and axial, radial and mean diffusivities) are not fibre-specific, which leads to complicated and potentially even erroneous interpretations of DTI findings in regions containing crossing fibres.^6^ In contrast, the recently developed fixel-based analysis (FBA) framework is based on a model of multiple fibre orientations within voxels (constrained spherical deconvolution).^7^ Because of this, FBA is able to characterise microstructural properties of each ’fixel’ (specific fibre population in any direction within an image voxel).^8^ In addition, FBA characterises morphological (macrostructural) properties of specific fibre populations. The FBA framework enables statistical analysis of the following parameters at every fixel across the whole-brain white matter: fibre density (FD), a microstructural measure of changes in the density of axons within a fibre population; fibre cross-section (FC), a macrostructural measure of changes in the cross-sectional area that a fibre population occupies, and; fibre density and cross-section (FDC), a combined measure of both of these microstructural and macrostructural properties.^8^ Decreases in FD can indicate axonal loss, whereas decreases in FC can indicate macroscopic fibre atrophy in diseases such as Alzheimer’s disease and multiple sclerosis.^9-11^

Previous studies have clearly documented the impact of preterm birth and associated perinatal risk factors on FBA metrics during the neonatal period.^12,13^ Infants born <31 weeks’ gestation and scanned at 38-44 weeks’ postmenstrual age had lower FD, FC and FDC in the corpus callosum, fornix, anterior commissure, optic radiation, cerebral and cerebellar peduncles, cingulum and superior longitudinal fasciculus compared with full-term (FT, born 38-41 weeks’ gestation) infants.^12^ In the same study, brain abnormality scored using the neonatal MRI scoring system of Kidokoro et al.^14^ was associated with reduced FD in the corpus callosum.^12^ In another study, earlier gestational age at birth and lower birth weight were associated with lower FD, FC and FDC in a number of white matter fibre tracts in infants born <33 weeks’ gestation and scanned at 38–47 weeks’ postmenstrual age.^13^

However, there have been no published applications of FBA to preterm populations beyond the neonatal period. Furthermore, there have been few longitudinal studies of white matter development during childhood and adolescence following preterm birth compared with FT birth; none using FBA and very few using DTI.^15,16^ As such, there is a need for long-term, longitudinal studies using advanced analysis methods to expand knowledge on the effects of VP birth on white matter development. In the current study, we apply the recently developed FBA framework to investigate the long-term implications of VP birth and associated perinatal risk factors on fibre microstructure and macrostructure over childhood and adolescence.

## Methods

### Participants

A total of 224 infants born VP (specifically, <30 weeks’ gestation or <1250 g) without genetic or congenital abnormalities were recruited between July 2001 and December 2003 from the Royal Women’s Hospital, Melbourne, into a prospective longitudinal cohort study. A total of 76 infants born FT (≥37 weeks’ gestation) were also recruited; 45 at birth and 31 at 2 years of age. The participants were invited to return for MRI at ages 7 and 13 years. Exclusions during image processing due to incomplete or incorrect image acquisitions or movement artefact resulted in the final sample shown in Supplementary Figure 1. The study was approved by the Human Research and Ethics Committees of the Royal Women’s Hospital and the Royal Children’s Hospital, Melbourne. Parents gave written informed consent for their child to participate.

### Perinatal data

Perinatal data included gestational age at birth, sex, and birth weight standardised for gestational age and sex (birth weight z-score).^17^ Major neonatal brain injuries (intraventricular haemorrhage^18^ and cystic periventricular leukomalacia) were diagnosed from serial cranial ultrasounds prior to term in the VP group. Additionally, the presence and severity of brain abnormalities were assessed from MRI at term-equivalent age using a semi-qualitative scoring system.^14^ A global brain abnormality score was calculated by summing abnormality scores across the white matter, cortical and deep grey matter, and cerebellum. Higher global brain abnormality scores indicate more severe brain abnormalities.

### MRI acquisition

At both the 7-year and 13-year follow-ups, MRI was conducted using a 3T scanner (Siemens Tim Trio, Erlangen, Germany).

At the 7-year follow-up, images acquired included *T*_1_-weighted images (3D ultrafast magnetization-prepared rapid gradient-echo (MP-RAGE) sequence, flip angle = 9°, repetition time = 1900 ms, echo time = 2.27 ms, field of view = 210 × 210 mm, matrix size = 256 × 256, echo spacing = 6.9 ms, voxel size = 0.8 mm isotropic, and acquisition time = 4 min 33 sec) and diffusion-weighted images (twice-refocused echo planar imaging sequence, 45 gradient directions at *b*-value = 3000 s/mm^2^, six *b* = 0 s/mm^2^ volumes, repetition time = 7400 ms, echo time = 106 ms, field of view = 240 × 240 mm, matrix size = 104 × 104, voxel size = 2.3 mm isotropic, and acquisition time = 6 min 52 sec).

At the 13-year follow-up, images acquired included *T*_1_-weighted images (3D multi-echo MP-RAGE sequence with prospective motion correction, repetition time 2530 = ms, echo times = 1.77, 3.51, 5.32 and 7.2 ms, flip angle = 7°, field of view = 230 × 209 mm, matrix = 256 × 230, interpolated = 256 × 256, voxel size = 0.9 mm isotropic, and acquisition time = 6 min 52 sec) and diffusion-weighted images (multi-band accelerated echo planar imaging pulse sequence, 60 gradient directions at *b*-value = 2800 s/mm^2^, four *b =* 0 s/mm^2^ images, repetition time = 3200 ms, echo time = 110 ms, field of view = 260 × 260 mm, matrix size = 110 × 110, voxel size = 2.4 mm isotropic, multi-band acceleration factor = 3, and acquisition time = 3 min 57 sec). A pair of *b* = 0 s/mm^2^ images with reversed phase encoding (anterior-posterior and posterior-anterior) were also acquired.

### Diffusion image pre-processing

Diffusion images were brain extracted using the Brain Extraction Tool (BET) from the Functional MRI of the Brain (FMRIB) Software Library (FSL; version 5.0.11).^19^ Brain masks were manually checked and edited as required. 7-year images were corrected for: (1) movement and eddy current-induced distortions [the FSL ‘eddy’ tool with slice-to-volume motion correction, outlier detection and replacement and *b*-vector reorientation^20-23^] and; (2) susceptibility-induced distortions [based on information from the *T*_1_-weighted images using BrainSuite version 18^24^]. 13-year images were corrected for: (1) susceptibility-induced distortions [based on information from the reversed phase-encoded images using the FSL ‘topup’ tool^25^] and; (2) motion and eddy-current-induced distortions [the FSL ‘eddy’ tool with slice-to-volume motion correction, outlier detection and replacement, and *b*-vector reorientation^20-23^]. We visually examined corrected diffusion images, and also obtained quantitative metrics relating to image quality using FSL’s QUality Assessment for DMRI (QUAD) and Study-wise QUality Assessment for DMRI (SQUAD) tools;^26^ this information resulted in exclusion of four images from the 7-year follow-up and one image from the 13-year follow-up. For the remaining images that were suitable for inclusion, quality control metrics generally did not differ between the VP and FT groups, but many metrics were poorer for the 7-year images compared with the 13-year images (Supplementary Table 1).

### Fixel-based analysis

Following pre-processing, images were analysed using recommended steps for a single-shell, single-tissue FBA^8^ in MRtrix3 (version 3.0_RC3).^27^ Images were corrected for bias fields.^28^ Global intensity normalisation was performed based on all images. Images were then upsampled to 1.3 mm^3^. Constrained spherical deconvolution was performed on all images using an average response function made from all images,^29^ resulting in fibre orientation distributions (FOD).

30 participants (15 VP and 15 FT) were randomly selected from each follow-up to generate a 7-year and a 13-year study-specific FOD template for the cross-sectional analyses. The total 60 images were used to generate a template for longitudinal analyses. Participants with major neonatal brain injuries were excluded from the template subset, consistent with previous studies.^13^ Age and sex ratio in the template subset did not differ to that of the remaining participants (all *p* > 0.1).

7-year and 13-year FOD images were registered to their respective FOD template, and all FOD images were registered to the longitudinal FOD template. We visually examined registrations, and excluded one 7-year VP image that had poor registration as a result of brain abnormalities. In all three template spaces, fixels were segmented and FBA metrics (FD, FC in log form, and FDC) were calculated as previously described.^8^ Whole brain tractography was performed on the FOD templates, with spherical-deconvolution informed filtering of tractograms (SIFT),^30^ which is required for statistical analysis using the connectivity-based fixel enhancement (CFE) method.^31^

### Statistical analysis

Statistical analyses were performed at each fixel across the whole-brain white matter using a General Linear Model. At each fixel, we compared FD, FC and FDC between the VP and FT groups separately at ages 7 and 13 years. These cross-sectional analyses were performed in the space of the respective cross-sectional template. All comparisons were adjusted for age and sex, and comparisons of FC and FDC were repeated adjusting for intracranial volume (generated from the *T*_1_)-weighted images using Statistical Parametric Mapping version 12), consistent with previous studies.^8,12,13^ Connectivity-based smoothing and statistical inference were performed using CFE.^31^ Non-parametric permutation testing (5000 permutations) was used to generate a family-wise error rate-corrected *p*-value for every individual fixel.

Again, using a General Linear Model, CFE and non-parametric permutation testing, we compared longitudinal changes in FD, FC and FDC at each fixel from age 7 to 13 years between the VP and FT groups, adjusting for change in age, sex and change in intracranial volume. To enable these longitudinal analyses, we subtracted each participant’s 7-year FD, FC and FDC image from their 13-year FD, FC and FDC image, and statistical comparisons were performed on the difference in FD, FC and FDC between ages. Furthermore, a modified version of CFE was used to enable the longitudinal analyses, as previously detailed.^32^ These longitudinal analyses were performed in the space of the longitudinal template.

As a secondary analysis, we repeated the above cross-sectional and longitudinal comparisons of FBA metrics at each fixel between the VP and FT groups excluding the subset of VP children who had intraventricular haemorrhage grade 3 or 4 and/or cystic periventricular leukomalacia, to ensure any group differences in FBA metrics were not driven by this subset.

Finally, using the same methods described above, in the VP group, we examined associations between perinatal data (gestational age at birth, birth weight z-score, sex, and neonatal global brain abnormality score) and: (1) FD, FC and FDC at each fixel at ages 7 and 13 years, and; (2) longitudinal changes in FD, FC and FDC at each fixel between ages 7 and 13 years. Again, cross-sectional analyses were performed in the space of the cross-sectional templates and longitudinal analyses in the space of the longitudinal templates.

Fixels were considered statistically significant at *p*<0.05, family-wise error rate-corrected. Significant fixels were anatomically localised by visual inspection and comparison against a white matter atlas.^33^ The results section focuses on major results that were confirmed in secondary analyses.

### Data availability statement

The data analysed during the current study are available from the corresponding author on reasonable request and completion of a data sharing agreement. The data are not publicly available due to ethical restrictions.

## Results

### Participant characteristics

The proportion of males and females did not differ significantly between the VP and FT groups (Table 1). The rate of major neonatal brain injuries was relatively low in the VP group. Neonatal global brain abnormality score was higher in the VP group compared with the FT group. Age at each MRI was similar for the VP and FT groups. Intracranial volume was smaller in the VP group than the FT group at ages 7 and 13 years.

**Table 1.** Characteristics of the participants included in the fixel-based analysis, separated by age (7 years and 13 years) and group (very preterm (VP) and full-term (FT)).

| Characteristic | Age 7 years |  |  | Age 13 years |  |  |
| --- | --- | --- | --- | --- | --- | --- |
|  | VP,<br>*n* = 138 | FT,<br>*n* = 32 | *p*-value<br>(VP-FT) | VP,<br>*n* = 130 | FT,<br>*n* = 45 | *p*-value<br>(VP-FT) |
| Gestational age at birth (weeks), mean (SD) | 27.5 (1.9) | 38.9 (1.3) | NA | 27.4 (1.9) | 39.0 (1.4) | NA |
| Birth weight (grams), mean (SD) | 977 (224) | 3247 (509) | NA | 964 (230) | 3316 (552) | NA |
| Females, *n* (%) | 71 (51) | 17 (53) | 0.86 | 61 (47) | 23 (51) | 0.63 |
| Intraventricular haemorrhage grade 3 or 4, *n* (%) | 5 (4) | NA | NA | 7 (5) | NA | NA |
| Cystic periventricular leukomalacia, *n* (%) | 4 (3) | NA | NA | 4 (3) | NA | NA |
| Neonatal global brain abnormality score, mean (SD) | 5.07 (2.89)<sup>a</sup> | 1.72 (1.44) | < 0.001 | 5.25 (3.00) | 1.50 (1.41)<sup>d</sup> | < 0.001 |
| Age at MRI (years), mean (SD) | 7.5 (0.2) | 7.6 (0.2) | 0.17 | 13.3 (0.4) | 13.3 (0.5) | 0.97 |
| Intracranial volume (cm<sup>3</sup>), mean (SD) | 1339 (114)<sup>b</sup> | 1432 (102)<sup>c</sup> | < 0.001 | 1441 (132) | 1526 (157) | < 0.001 |

<sup>a</sup>*n* = 137; <sup>b</sup>*n* = 135; <sup>c</sup>*n* = 30 (there were 5 participants at the 7-year follow-up who
were included in the fixel-based analysis but were missing intracranial volume data,
because their *T*<sub>1</sub>-weighted images had movement artefact and/or tissue
segmentation failed for these participants, despite them having good quality diffusion-weighted images); <sup>d</sup> $n = 24$ .

Characteristics of the included participants were generally similar to those of the non-participants (the participants, of the original *n* = 224 VP and *n* = 76 FT participants, who were recruited into the study but could not be followed up or were excluded from the current analysis). However, included VP participants had lower neonatal global brain abnormality scores than VP non-participants (for the 7-year follow-up sample, mean (SD)= 5 (3) versus 7 (4), *p* = 0.001; for the 13-year follow-up sample, mean (SD)= 5 (3) versus 6 (4), *p* = 0.03).

### Group differences

VP children had significantly lower FD, FC and FDC than FT children in many fibre tracts at ages 7 and 13 years (Figure 1). At age 7 years, FD was 21% lower on average across all significantly affected fibre tracts in VP children compared with FT children, and was up to 54% lower in some white matter tracts (such as the tapetum and fornix). FC group differences were smaller in magnitude, but more widespread across the white matter, than the FD differences. A similar pattern was observed at age 13 years; FD was on average 16% lower in the VP group compared with the FT group, and up to 51% lower in tracts such as the tapetum and fornix, and again FC differences were smaller in magnitude but more widespread than the FD differences. Additionally, more fibre tracts differed between groups at age 13 years than 7 years; for example, 7% versus 13% of the white matter at ages 7 and 13 years respectively had lower FC in the VP group compared with the FT group. The FC and FDC group differences at both ages were no longer significant after adjusting for intracranial volume.

**Figure 1.**
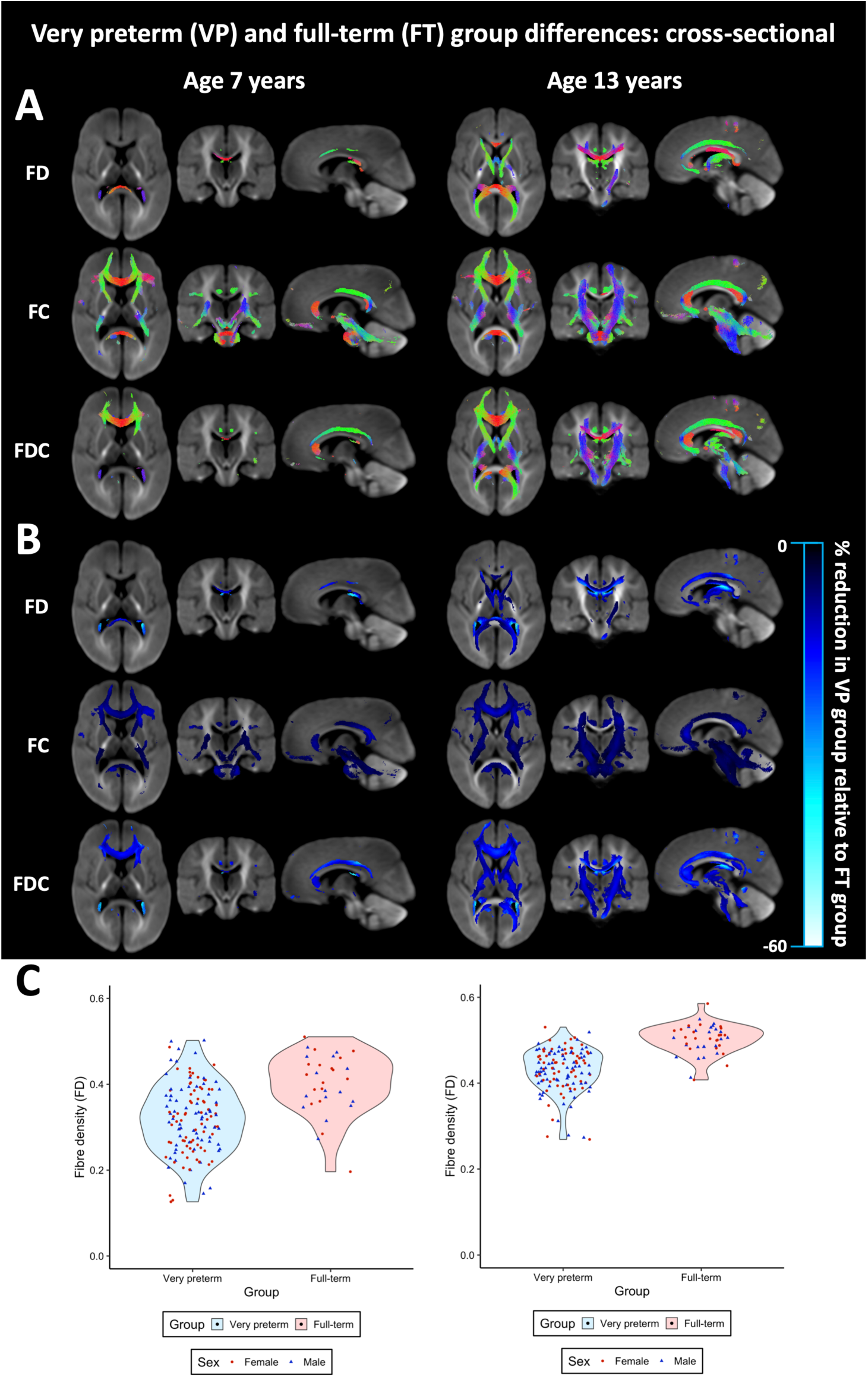
Cross-sectional group differences in fixel-based analysis metrics. The fibre tracts that had significantly lower fibre density (FD), fibre cross-section (FC), and fibre density and cross-section (FDC) in the very preterm (VP) group compared with the full-term (FT) group cross-sectionally at ages 7 and 13 years (*p* < 0.05, family-wise error rate-corrected). Fibre tracts are coloured by (A) direction (anterior-posterior = green; superior-inferior = blue; left-right = red) and (B) the percentage reduction in FD, FC or FDC in the VP group compared with the FT group. In (C), FD values at age 7 (left) and 13 (right) years, averaged across the significant tracts in (A) and (B), are shown as violin plots. All significant regions presented were obtained from models adjusted for age and sex. For FC and FDC, there were no significant regions after additionally adjusting for intracranial volume.

Between ages 7 and 13 years, FDC in the splenium of the corpus callosum, right cerebral peduncle and right posterior limb of the internal capsule increased significantly less in VP children compared with FT children, even after adjusting for intracranial volume (Figure 2). In these fibre tracts, the increase over time was on average 26% less in the VP group compared with the FT group.

**Figure 2.**
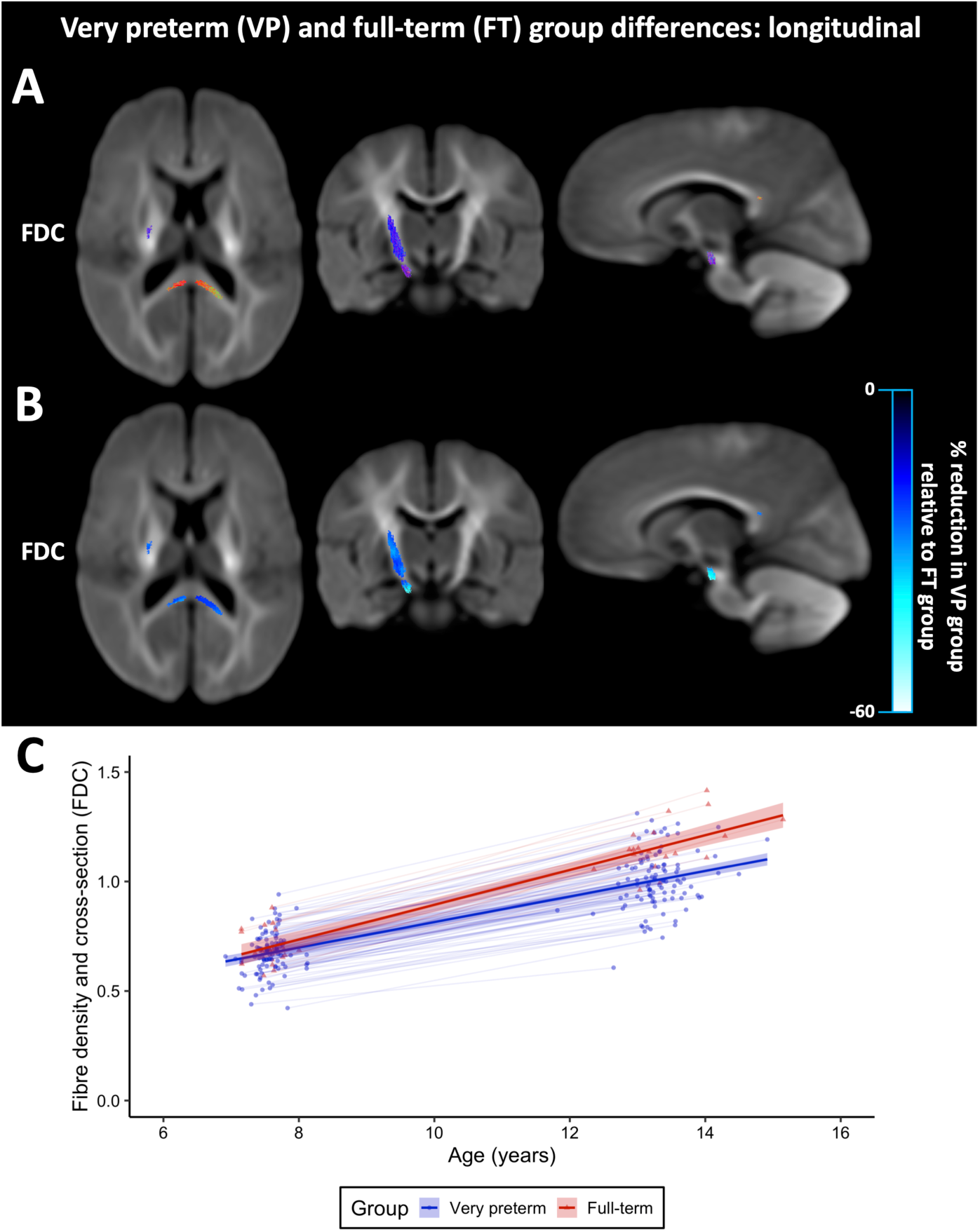
Longitudinal group differences in fixel-based analysis metrics. The fibre tracts in which fibre density and cross-section (FDC) increased significantly less between ages 7 and 13 years in the very preterm (VP) group compared with the full-term (FT) group (*p* < 0.05, family-wise error rate-corrected). Fibre tracts are coloured by (A) direction (anterior-posterior = green; superior-inferior = blue; left-right = red) and (B) the percentage by which the VP group increased less over time compared with the FT group. In (C), FDC values averaged across the significant tracts in (A) and (B) have been plotted against age. All significant regions presented were obtained from models adjusted for change in age, sex and change in intracranial volume.

### Perinatal factors

Earlier gestational age at birth was significantly associated with lower FD, FC and FDC in several fibre tracts, particularly the corpus callosum, tapetum and anterior commissure, at ages 7 and 13 years. Lower birth weight z-score was significantly associated with lower FC and FDC in many fibre tracts throughout the white matter at both ages. For both gestational age at birth and birth weight z-score, associations with FC and FDC were not significant after adjusting for intracranial volume (Figure 3).

**Figure 3.**
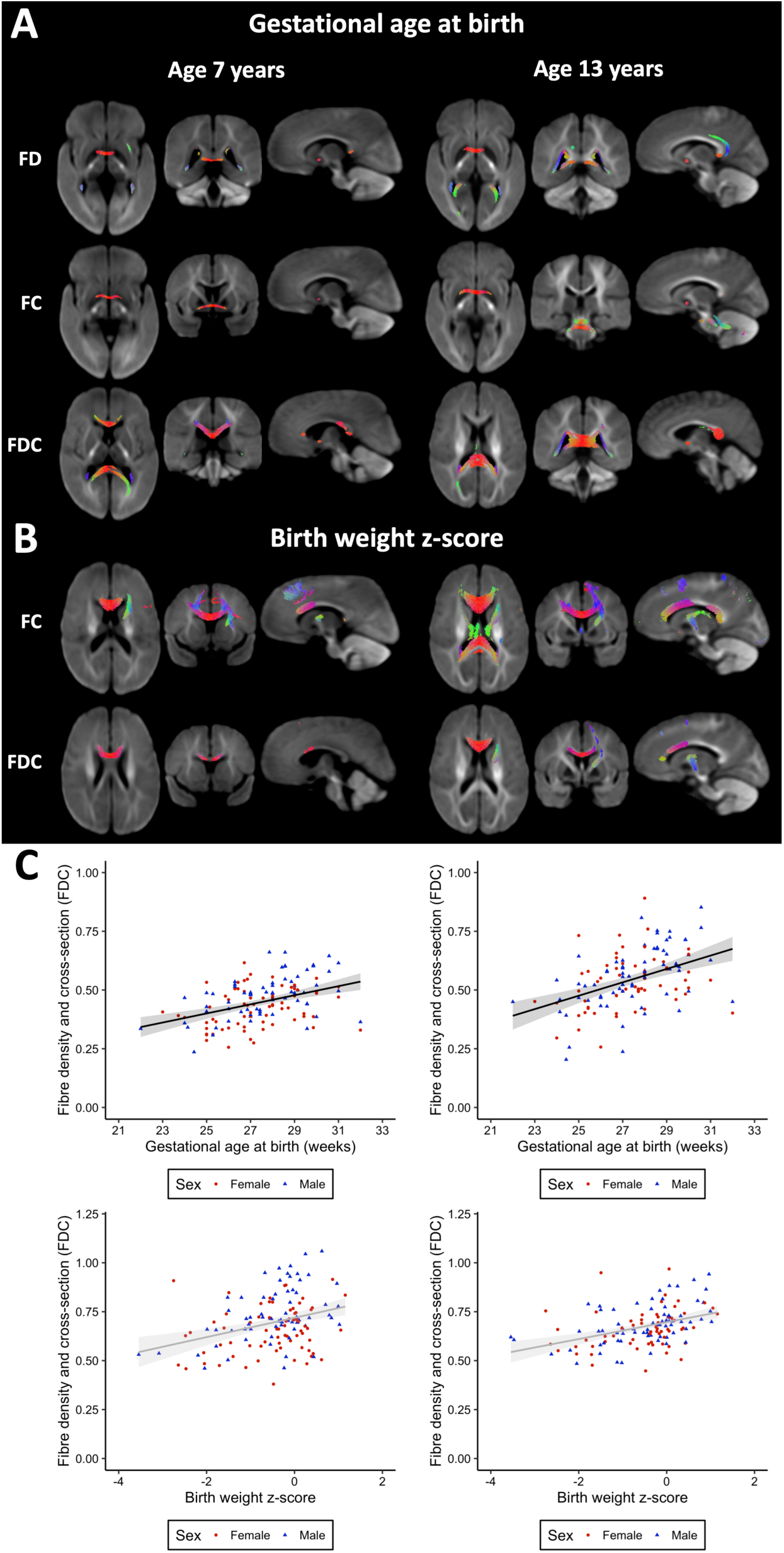
Associations between gestational age at birth, birth weight z-score, and fixel-based analysis metrics. The fibre tracts in which earlier gestational age at birth (A) and lower birth weight z-score (B) were significantly associated with lower fibre density (FD), fibre cross-section (FC) and fibre density and cross-section (FDC) cross-sectionally at ages 7 and 13 years in the very preterm (VP) group (*p* < 0.05, family-wise error rate-corrected). Fibre tracts are coloured by direction (anterior-posterior = green; superior-inferior = blue; left-right = red). In (C), FDC values at age 7 (left) and 13 (right) years, averaged across the significant tracts in (A) and (B), have been plotted against gestational age at birth and birth weight z-score. All significant regions presented were obtained from models adjusted for age and sex. For FC and FDC, there were no significant regions after additionally adjusting for intracranial volume.

VP males had significantly larger FC and FDC throughout the white matter compared with VP females at ages 7 and 13 years, although these differences were no longer significant after adjusting for intracranial volume. Between ages 7 and 13 years, VP females had a significantly greater increase in FC in the corpus callosum than VP males, even after adjusting for intracranial volume (Figure 4).

**Figure 4.**
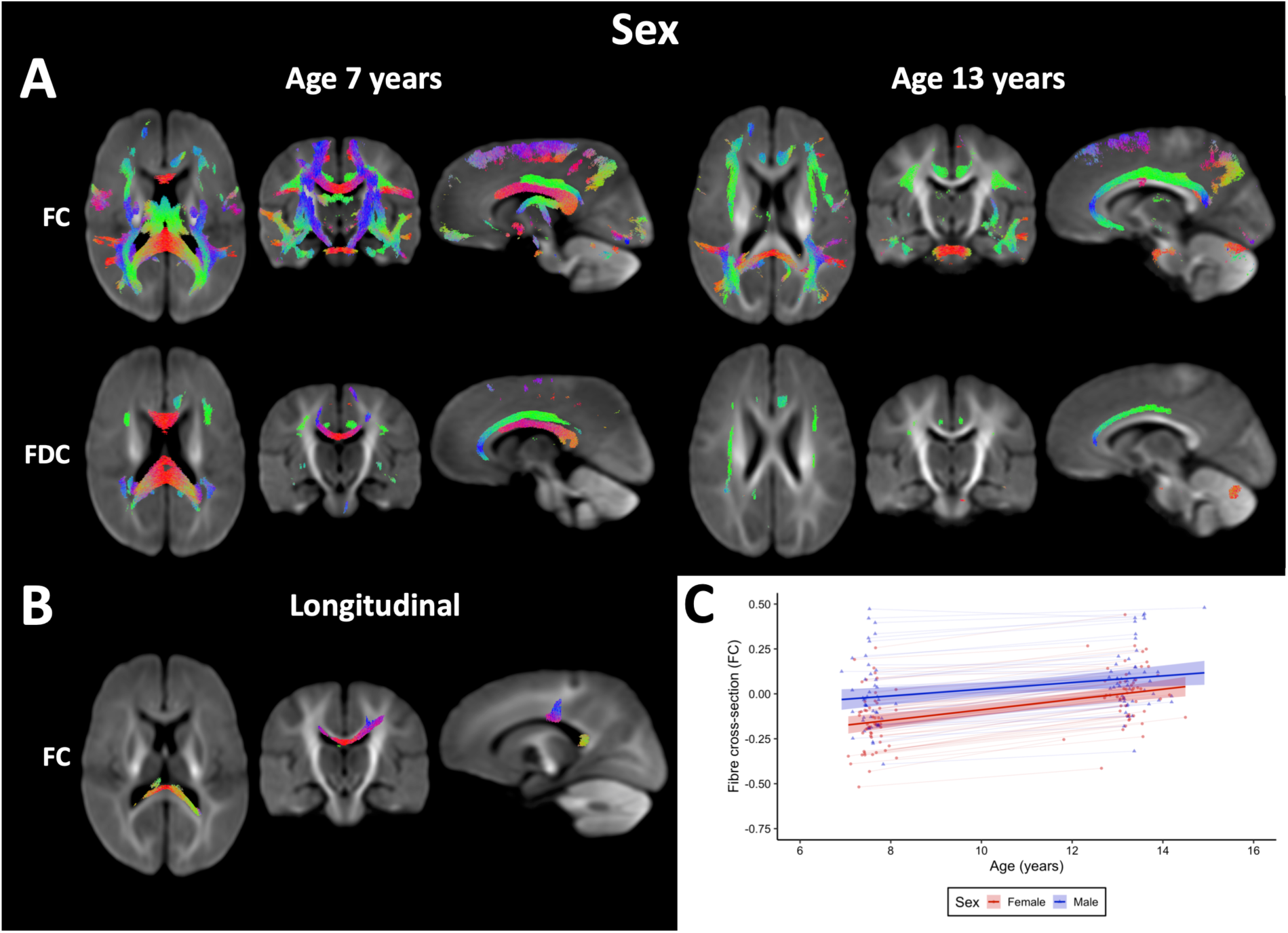
Differences in fixel-based analysis (FBA) metrics between very preterm (VP) males and VP females.

The fibre tracts in which (A) fibre cross-section (FC) and fibre density and cross-section (FDC) were significantly higher in VP males compared with VP females at ages 7 and 13 years, and (B) FC increased significantly more over time in VP females compared with VP males (*p* < 0.05, family-wise error rate-corrected). Fibre tracts are coloured by direction (anterior-posterior = green; superior-inferior = blue; left-right = red). In (C), FC values averaged across the significant tracts in (B) have been plotted against age. In part A, all significant regions presented were obtained from models adjusted for age; for FC and FDC, there were no significant regions after additionally adjusting for intracranial volume. In part B, all significant regions presented were obtained from models adjusted for change in age and change in intracranial volume.

Higher neonatal global brain abnormality scores were significantly associated with lower FD, FC and FDC throughout the white matter at ages 7 and 13 years, although the associations with FC and FDC were no longer significant after adjusting for intracranial volume. Higher neonatal global brain abnormality scores were also significantly associated with less increase in FDC in the corpus callosum between ages 7 and 13 years, even after adjusting for intracranial volume (Figure 5).

**Figure 5.**
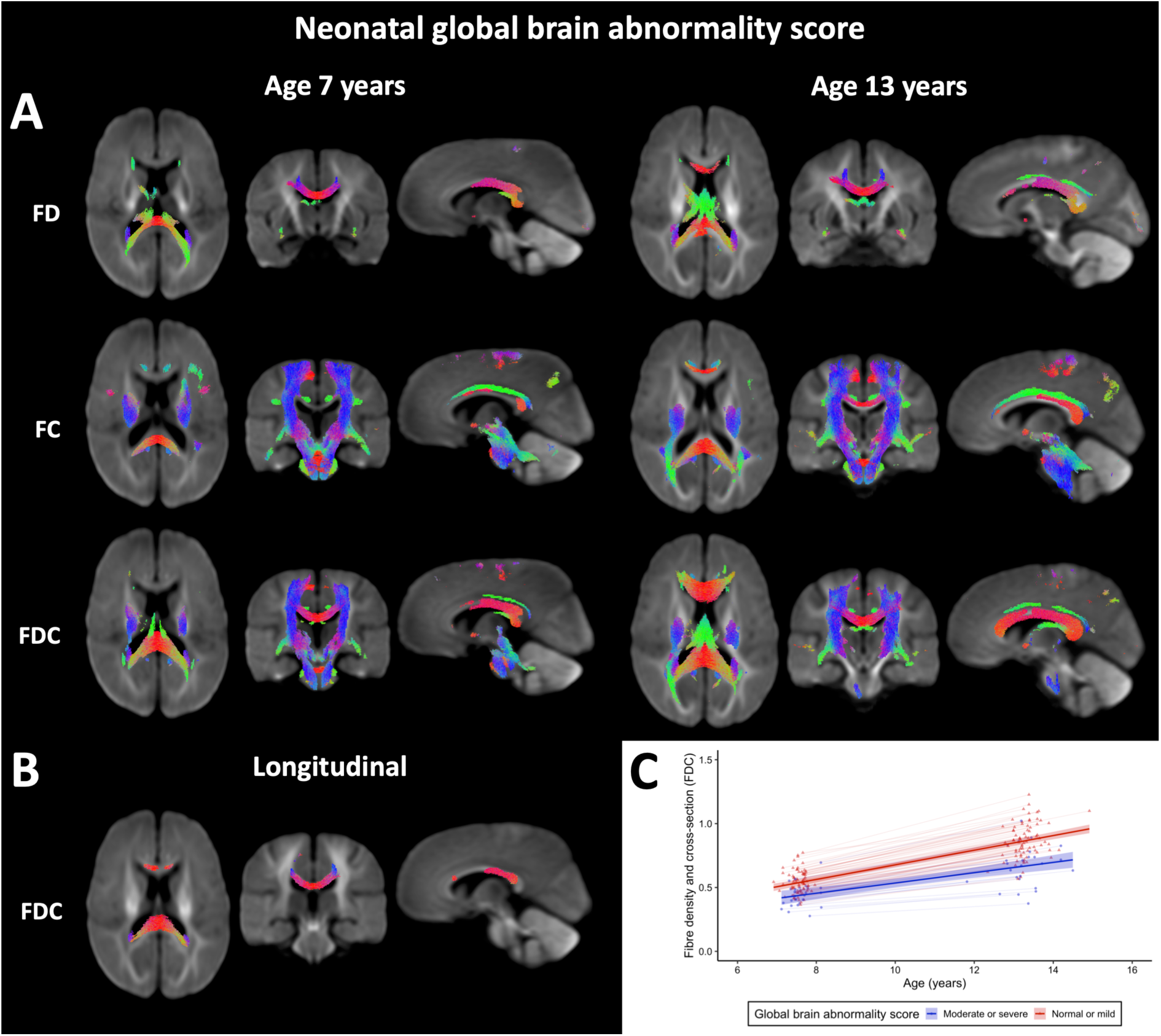
Associations between neonatal brain abnormality score and fixel-based analysis metrics. The fibre tracts in which higher neonatal global brain abnormality scores were significantly associated with (A) lower fibre density (FD), fibre cross-section (FC) and fibre density and cross-section (FDC) at ages 7 and 13 years, and (B) less increase in FDC between ages 7 and 13 years, in the very preterm (VP) group (*p* < 0.05, family-wise error rate-corrected). Fibre tracts are coloured by direction (anterior-posterior = green; superior-inferior = blue; left-right = red). In (C), FDC values averaged across the significant tracts in (B) have been plotted against age (separated for VP children with normal/mild and moderate/severe neonatal global brain abnormality scores). In part A, all significant regions presented were obtained from models adjusted for age and sex; for FC and FDC, there were no significant regions after additionally adjusting for intracranial volume. In part B, all significant regions presented were obtained from models adjusted for change in age, sex and change in intracranial volume.

## Discussion

We have presented a unique long-term, longitudinal study of white matter development up to 13 years after VP birth in comparison with FT birth. Using the recently developed FBA framework, we revealed fibre tract-specific findings: (1) VP children have axonal reductions across many fibre tracts at ages 7 and 13 years compared with FT children; (2) VP children have slower axonal growth over time specifically in the corpus callosum and corticospinal tract, and; (3) earlier gestational age at birth, lower birth weight, and neonatal brain abnormalities are important perinatal factors associated with later axonal reductions within the VP population.

At high diffusion weightings (*b*-values ≥ 3000 s/mm^2^), the extra-axonal water signal is strongly attenuated, leaving the remaining signal localised to the intra-axonal water compartment, and allowing us to make fibre-specific inferences using FBA.^8,34^ The observed lower FD in VP children compared with FT children suggests VP children have lower axonal densities within fibre tracts, which could reflect axon loss or smaller axonal diameters.^35^ In addition to microstructural differences within fibre tracts, the lower FC in VP children compared with FT children suggests VP children also have lower total cross-sectional area of fibre tracts. Additionally, lower myelination could increase the exchange of water between the intra-axonal and extra-axonal spaces, resulting in an apparent decrease in the volume of the intra-axonal compartment, and hence a decrease in FD.^12,35^ Therefore, the current results could also indicate that VP children have less myelination than FT children. These findings suggest there is a decreased capacity to transfer information between brain regions in VP children compared with FT children.^8^

VP children had lower FD in many specific fibre pathways compared with FT children. The fibre tracts predominantly affected included the corpus callosum, tapetum, inferior fronto-occipital fasciculus, fornix and cingulum at both ages, as well as the corticospinal tract and anterior limb of the internal capsule at age 13 years. These fibre tracts are rapidly developing during childhood,^36^ suggesting VP children have axonal reductions in developmentally sensitive regions. The group differences in FC and FDC were located in similar but more widespread tracts. However, they were no longer significant after adjusting for intracranial volume, indicating that macrostructural axonal reductions in VP children compared with FT children are commensurate with the known smaller head size in VP children.^37^ Our findings are consistent with previous studies at term-equivalent age,^12^ suggesting that early axonal reductions are still present in childhood and adolescence.

In the premature brain an initial insult such as hypoxia, ischemia or inflammation is thought to lead to injury of pre-myelinating oligodendrocytes, followed by impaired myelination and axonal growth.^3^ Recently, lower FC measured using FBA in the periventricular white matter, hippocampus and cerebellar white matter was related to histological markers of hypomyelination and axonal disorganisation in a growth-restricted, preterm animal model.^38^ The lower FC and histological abnormalities were found immediately after preterm birth, indicating the white matter injury occurred antenatally.^38^ Our study provides *in vivo* markers of the long-term effects of possible underlying antenatal white matter injury and/or altered development in VP children.

Longitudinally, VP children had slower FDC development in the splenium of the corpus callosum and corticospinal tract between ages 7 and 13 years than FT children. Previous diffusion MRI studies have shown that typical white matter development is most rapid in the fetal and early postnatal period, but there is ongoing white matter development over childhood and adolescence in the form of increasing axonal density and myelination.^32,36,39-41^ In the corpus callosum (whose development was affected in our VP sample) the number of axons is typically close to a maximum at birth after which no new axons are formed, but structural changes including axonal pruning, redirection and myelination occur, which are more pronounced in the posterior (splenium) than anterior (genu) subdivisions.^41,42^ Therefore, our results may reflect reduced axonal growth (less increase in axonal diameter) and/or reduced myelination specifically in the splenium and corticospinal tract between ages 7 and 13 years in VP children compared with FT children.

Earlier gestational age at birth, lower birth weight z-score and neonatal brain abnormalities were related to lower FD, FC and/or FDC in many specific fibre pathways at both ages; neonatal brain abnormalities were additionally related to slower change in FDC in the corpus callosum over time. This agrees with previous findings at term-equivalent age.^12,13^ We also found VP males had larger FC and FDC throughout the white matter than VP females, proportional to their larger intracranial volume. Equivalent sex-based differences in FBA metrics were found previously in the neonatal period.^13^ In our study, we additionally found that VP females had faster axonal growth in the corpus callosum than VP males. This could reflect the developmental vulnerability of VP males that has been previously documented,^43^ or influences of puberty.^32,44^

This study is strengthened by the large cohort followed up over a long period of time. Some participants could not be followed up, however included participants were generally representative of the originally recruited sample, except they had lower neonatal brain abnormality scores. This suggests results may have been even more pronounced had all of the recruited participants been included. We acknowledge there were differences in MRI sequences and image quality between the 7-year and 13-year follow-ups. However, this should not influence the strength or magnitude of cross-sectional and longitudinal group differences, because image sequences and quality were consistent between the VP and FT groups.

## Conclusions

Using a long-term, longitudinal study design combined with advanced diffusion MRI, we found that VP birth and concomitant perinatal risk factors are associated with fibre-specific disruptions to axonal development, up to 13 years after birth. Understanding the specific white matter fibre properties such as density and cross-section affected by VP birth may in future enable identification of at-risk children and development of targeted interventions to improve outcomes for the vulnerable VP population.

## Acknowledgements

This research was conducted within the Victorian Infant Brain Study (VIBeS) and Developmental Imaging research groups, Murdoch Children’s Research Institute and the Children’s MRI Centre, the Royal Children’s Hospital, Melbourne, Victoria. We thank members of the VIBeS and Developmental Imaging teams, the Royal Children’s Hospital Medical Imaging staff for their assistance and expertise in the collection of the MRI data included in this study, and the children and families who participated. This research was supported by the Australian National Health and Medical Research Council (NHMRC; Centre for Research Excellence 546519, 1060733 and 1153176; Project Grant 237117, 491209 and 1066555; Senior Research Fellowship 1081288 to PJA; Investigator Grant 1176077 to PJA; Career Development Fellowship 1085754 to DKT and 1141354 to JLYC, and; Early Career Fellowship 1012236 to DKT), the Murdoch Children’s Research Institute, the Royal Children’s Hospital, the Royal Children’s Hospital Foundation (RCH1000 to JYMY), the Department of Paediatrics at the University of Melbourne, and the Victorian Government’s Operational Infrastructure Support Program. The funding organisations/sponsors had no role in the design and conduct of the study; collection, management, analysis, and interpretation of the data; preparation, review, or approval of the manuscript; and decision to submit the manuscript for publication. The authors have no potential conflicts of interest to disclose.

## Supplementary Material

**Supplementary Figure 1.**
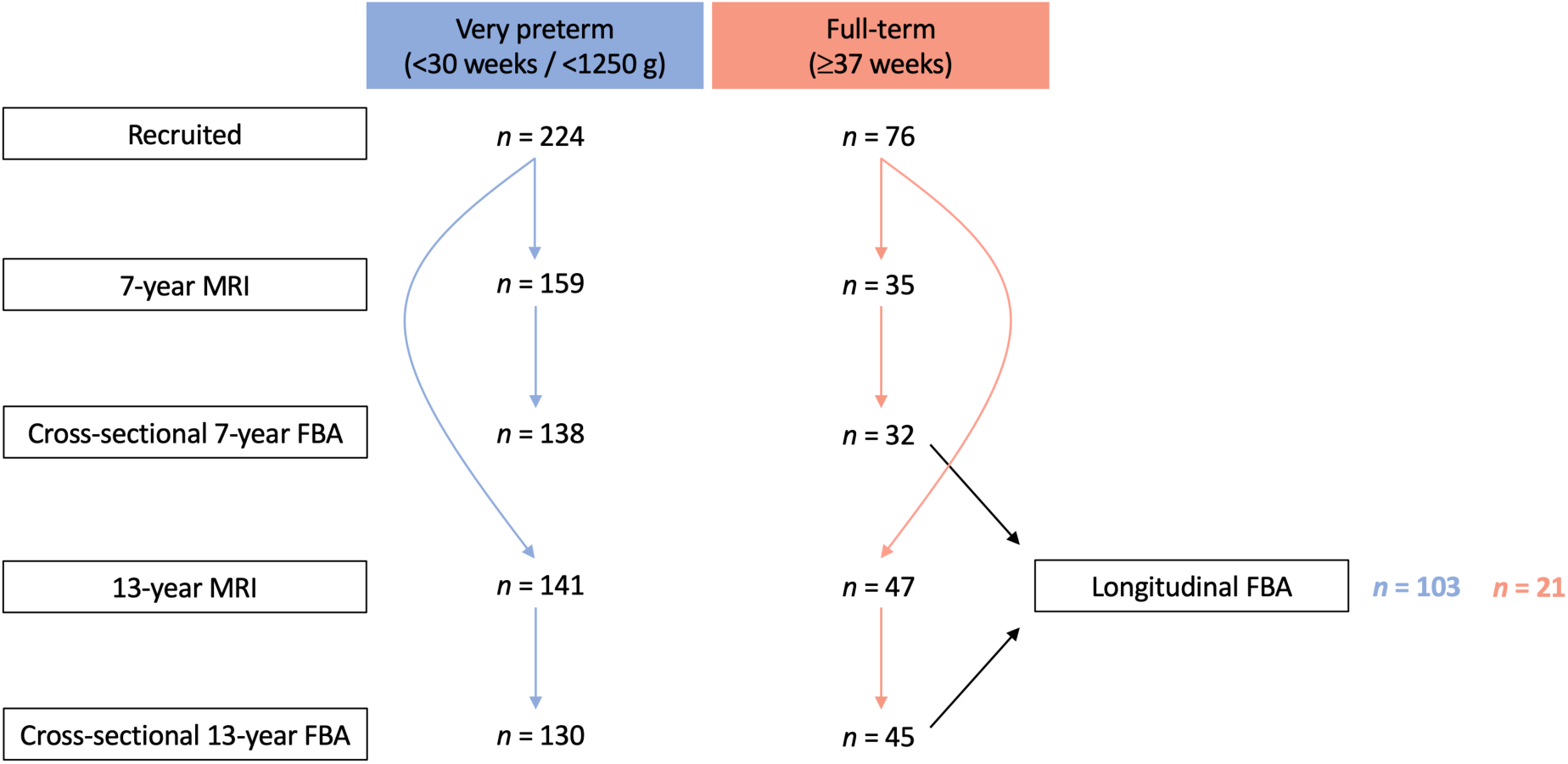
Participant flowchart. The number of very preterm and full-term children who were recruited, completed magnetic resonance imaging (MRI), and were included in the fixel-based analyses (FBA) at ages 7 and 13 years. The longitudinal FBA was based on children who had usable images at both ages.

**Supplementary Table 1.** **Quality control metrics for the images of the participants included in the fixel-based analysis, presented separately for the 7-year and 13-year images, and separately for the very preterm (VP) and full-term (FT) groups.**

| Quality control metric | 7-year |  |  |  |  | 13-year |  |  |  |  | *p*-value (7-13) |
| --- | --- | --- | --- | --- | --- | --- | --- | --- | --- | --- | --- |
|  | VP mean | VP SD | FT mean | FT SD | *p*-value (VP-FT) | VP mean | VP SD | FT mean | FT SD | *p*-value (VP-FT) |  |
| Average absolute motion (mm) | 0.82 | 0.40 | 0.86 | 0.39 | 0.583 | 0.66 | 0.32 | 0.66 | 0.35 | 0.976 | <0.001 |
| Average relative motion (mm) | 0.40 | 0.19 | 0.45 | 0.26 | 0.248 | 0.45 | 0.29 | 0.48 | 0.26 | 0.581 | 0.196 |
| Average translations (mm) |  |  |  |  |  |  |  |  |  |  |  |
| x | -0.08 | 0.23 | -0.10 | 0.23 | 0.603 | -0.004 | 0.18 | -0.001 | 0.14 | 0.922 | 0.002 |
| y | 0.18 | 0.33 | 0.06 | 0.30 | 0.080 | -0.16 | 0.25 | -0.19 | 0.32 | 0.545 | <0.001 |
| z | 0.07 | 0.37 | -0.06 | 0.29 | 0.068 | -0.22 | 0.28 | -0.19 | 0.29 | 0.581 | <0.001 |
| Average rotations (deg) |  |  |  |  |  |  |  |  |  |  |  |
| x | -0.004 | 0.41 | -0.07 | 0.43 | 0.381 | 0.08 | 0.20 | 0.06 | 0.16 | 0.569 | 0.093 |
| y | -0.03 | 0.24 | 0.04 | 0.22 | 0.151 | -0.003 | 0.16 | 0.02 | 0.15 | 0.396 | 0.216 |
| z | -0.12 | 0.35 | -0.08 | 0.44 | 0.551 | 0.08 | 0.27 | 0.12 | 0.25 | 0.397 | <0.001 |
| Average standard deviation translations (mm) |  |  |  |  |  |  |  |  |  |  |  |
| x | 0.04 | 0.02 | 0.04 | 0.02 | 0.569 | 0.02 | 0.01 | 0.02 | 0.01 | 0.498 | <0.001 |
| y | 0.08 | 0.04 | 0.08 | 0.04 | 0.739 | 0.06 | 0.02 | 0.06 | 0.02 | 0.955 | <0.001 |
| z | 0.08 | 0.09 | 0.07 | 0.06 | 0.698 | 0.05 | 0.03 | 0.04 | 0.02 | 0.021 | <0.001 |
| Average standard deviation rotations (deg) |  |  |  |  |  |  |  |  |  |  |  |
| x | 0.13 | 0.14 | 0.18 | 0.17 | 0.083 | 0.08 | 0.04 | 0.09 | 0.06 | 0.152 | <0.001 |
| y | 0.06 | 0.03 | 0.06 | 0.04 | 0.284 | 0.05 | 0.03 | 0.05 | 0.02 | 0.964 | 0.009 |
| z | 0.06 | 0.04 | 0.07 | 0.04 | 0.515 | 0.05 | 0.03 | 0.05 | 0.03 | 0.690 | 0.049 |
| Standard deviation eddy current linear terms (Hz/mm) |  |  |  |  |  |  |  |  |  |  |  |
| x | 0.02 | 0.01 | 0.02 | 0.01 | 0.710 | 0.02 | 0.01 | 0.02 | 0.01 | 0.813 | 0.820 |
| y | 0.03 | 0.01 | 0.03 | 0.01 | 0.878 | 0.02 | 0.01 | 0.02 | 0.01 | 0.441 | <0.001 |
| z | 0.02 | 0.01 | 0.03 | 0.01 | 0.055 | 0.02 | 0.01 | 0.02 | 0.01 | 0.964 | <0.001 |
| Susceptibility-induced distortions (standard deviation voxel displacement, mm) | NA | NA | NA | NA | NA | 0.91 | 0.16 | 0.97 | 0.14 | 0.043 | NA |
| Total number of outliers (%) | 1.70 | 1.05 | 2.26 | 1.53 | 0.015 | 0.79 | 0.41 | 0.76 | 0.45 | 0.729 | <0.001 |
| Average b0 signal-to-noise ratio | 29.54 | 4.23 | 29.00 | 3.93 | 0.508 | 34.10 | 3.93 | 34.89 | 3.92 | 0.245 | <0.001 |
| Average diffusion-weighted contrast-to-noise ratio | 2.07 | 0.65 | 2.25 | 0.77 | 0.186 | 2.08 | 0.52 | 2.10 | 0.45 | 0.796 | 0.262 |
| Average effective diffusion-weighted contrast-to-noise ratio | 1.22 | 0.38 | 1.32 | 0.45 | 0.186 | 1.20 | 0.30 | 1.21 | 0.26 | 0.796 | 0.117 |

Detailed information on the quality control metrics is provided in a previous publication.<sup>26</sup> The *p*-value for the 7-year compared with 13-year images is based on the 103 VP and 21 FT participants who had usable data at both follow-ups. 7-year images are NA for susceptibility-induced distortions because these were not corrected using the FSL ‘topup’ tool due to lack of reverse phase-encode images. Importantly, the effective contrast-to-noise ratio metric did not differ between 7-year and 13-year images (this metric takes differences in voxel sizes and number of acquired volumes into account, and therefore is a good metric for comparing image quality between studies, though it still depends on *b*-value, which differed slightly between the follow-up studies).

## References

1. Saigal S, Doyle LW. An overview of mortality and sequelae of preterm birth from infancy to adulthood. Lancet. 2008;371(9608):261–269.

2. Blencowe H, Cousens S, Oestergaard MZ, et al. National, regional, and worldwide estimates of preterm birth rates in the year 2010 with time trends since 1990 for selected countries: a systematic analysis and implications. Lancet. 2012;379(9832):2162–2172.

3. Volpe JJ. Dysmaturation of Premature Brain: Importance, Cellular Mechanisms, and Potential Interventions. Pediatr Neurol. 2019;95:42–66.

4. Barnett ML, Tusor N, Ball G, et al. Exploring the multiple-hit hypothesis of preterm white matter damage using diffusion MRI. Neuroimage Clin. 2018;17:596–606.

5. Jeurissen B, Leemans A, Tournier JD, Jones DK, Sijbers J. Investigating the prevalence of complex fiber configurations in white matter tissue with diffusion magnetic resonance imaging. Hum Brain Mapp. 2013;34(11):2747–2766.

6. Jones DK, Knosche TR, Turner R. White matter integrity, fiber count, and other fallacies: the do’s and don’ts of diffusion MRI. Neuroimage. 2013;73:239–254.

7. Tournier JD, Calamante F, Connelly A. Robust determination of the fibre orientation distribution in diffusion MRI: non-negativity constrained superresolved spherical deconvolution. Neuroimage. 2007;35(4):1459–1472.

8. Raffelt DA, Tournier JD, Smith RE, et al. Investigating white matter fibre density and morphology using fixel-based analysis. Neuroimage. 2017;144(Pt A):58–73.

9. Gajamange S, Raffelt D, Dhollander T, et al. Fibre-specific white matter changes in multiple sclerosis patients with optic neuritis. Neuroimage Clin. 2018;17:60–68.

10. Mito R, Raffelt D, Dhollander T, et al. Fibre-specific white matter reductions in Alzheimer’s disease and mild cognitive impairment. Brain. 2018;141(3):888–902.

11. Rojas-Vite G, Coronado-Leija R, Narvaez-Delgado O, et al. Histological validation of per-bundle water diffusion metrics within a region of fiber crossing following axonal degeneration. Neuroimage. 2019;201:116013.

12. Pannek K, Fripp J, George JM, et al. Fixel-based analysis reveals alterations is brain microstructure and macrostructure of preterm-born infants at term equivalent age. Neuroimage Clin. 2018;18:51–59.

13. Pecheva D, Tournier JD, Pietsch M, et al. Fixel-based analysis of the preterm brain: Disentangling bundle-specific white matter microstructural and macrostructural changes in relation to clinical risk factors. Neuroimage Clin. 2019;23:101820.

14. Kidokoro H, Neil JJ, Inder TE. New MR imaging assessment tool to define brain abnormalities in very preterm infants at term. Am J Neuroradiol. 2013;34(11):2208–2214.

15. Jo HM, Cho HK, Jang SH, et al. A comparison of microstructural maturational changes of the corpus callosum in preterm and full-term children: a diffusion tensor imaging study. Neuroradiology. 2012;54(9):997–1005.

16. Thompson DK, Lee KJ, van Bijnen L, et al. Accelerated corpus callosum development in prematurity predicts improved outcome. Hum Brain Mapp. 2015;36(10):3733–3748.

17. Cole TJ, Freeman JV, Preece MA. British 1990 growth reference centiles for weight, height, body mass index and head circumference fitted by maximum penalized likelihood. Stat Med. 1998;17(4):407–429.

18. Papile LA, Burstein J, Burstein R, Koffler H. Incidence and evolution of subependymal and intraventricular hemorrhage: a study of infants with birth weights less than 1,500 gm. J Pediatr. 1978;92(4):529–534.

19. Smith SM. Fast robust automated brain extraction. Hum Brain Mapp. 2002;17(3):143–155.

20. Andersson JLR, Sotiropoulos SN. An integrated approach to correction for off-resonance effects and subject movement in diffusion MR imaging. Neuroimage. 2016;125:1063–1078.

21. Andersson JLR, Graham MS, Drobnjak I, Zhang H, Filippini N, Bastiani M. Towards a comprehensive framework for movement and distortion correction of diffusion MR images: Within volume movement. Neuroimage. 2017;152:450–466.

22. Andersson JLR, Graham MS, Zsoldos E, Sotiropoulos SN. Incorporating outlier detection and replacement into a non-parametric framework for movement and distortion correction of diffusion MR images. Neuroimage. 2016;141:556–572.

23. Leemans A, Jones DK. The B-matrix must be rotated when correcting for subject motion in DTI data. Magn Reson Med. 2009;61(6):1336–1349.

24. Bhushan C, Haldar JP, Joshi AA, Leahy RM. Correcting Susceptibility-Induced Distortion in Diffusion-Weighted MRI using Constrained Nonrigid Registration. 2012 Asia-Pacific Signal and Information Processing Association Annual Summit and Conference (Apsipa Asc). 2012. <Go to ISI>://WOS:000319456200258.

25. Andersson JL, Skare S, Ashburner J. How to correct susceptibility distortions in spin-echo echo-planar images: application to diffusion tensor imaging. Neuroimage. 2003;20(2):870–888.

26. Bastiani M, Cottaar M, Fitzgibbon SP, et al. Automated quality control for within and between studies diffusion MRI data using a non-parametric framework for movement and distortion correction. Neuroimage. 2019;184:801–812.

27. Tournier JD, Smith R, Raffelt D, et al. MRtrix3: A fast, flexible and open software framework for medical image processing and visualisation. Neuroimage. 2019;202:116137.

28. Tustison NJ, Avants BB, Cook PA, et al. N4ITK: improved N3 bias correction. IEEE Trans Med Imaging. 2010;29(6):1310–1320.

29. Tournier JD, Calamante F, Connelly A. Determination of the appropriate b value and number of gradient directions for high-angular-resolution diffusion-weighted imaging. NMR Biomed. 2013;26(12):1775–1786.

30. Smith RE, Tournier JD, Calamante F, Connelly A. SIFT: Spherical-deconvolution informed filtering of tractograms. Neuroimage. 2013;67:298–312.

31. Raffelt DA, Smith RE, Ridgway GR, et al. Connectivity-based fixel enhancement: Whole-brain statistical analysis of diffusion MRI measures in the presence of crossing fibres. Neuroimage. 2015;117:40–55.

32. Genc S, Smith RE, Malpas CB, et al. Development of white matter fibre density and morphology over childhood: A longitudinal fixel-based analysis. Neuroimage. 2018;183:666–676.

33. Oishi K, Faria A, van Zijl PCM, Mori S. MRI atlas of human white matter-second edition. Elseiver B.V.; 2011.

34. Genc S, Tax CMW, Raven EP, Chamberland M, Parker GD, Jones DK. Impact of b-value on estimates of apparent fibre density. bioRxiv. 2020; https://doi.org/10.1101/2020.01.15.905802.

35. Raffelt D, Tournier JD, Rose S, et al. Apparent Fibre Density: a novel measure for the analysis of diffusion-weighted magnetic resonance images. Neuroimage. 2012;59(4):3976–3994.

36. Lebel C, Beaulieu C. Longitudinal development of human brain wiring continues from childhood into adulthood. J Neurosci. 2011;31(30):10937–10947.

37. Monson BB, Anderson PJ, Matthews LG, et al. Examination of the Pattern of Growth of Cerebral Tissue Volumes From Hospital Discharge to Early Childhood in Very Preterm Infants. JAMA Pediatr. 2016;170(8):772–779.

38. Malhotra A, Sepehrizadeh T, Dhollander T, et al. Advanced MRI analysis to detect white matter brain injury in growth restricted newborn lambs. Neuroimage Clin. 2019;24:101991.

39. Geeraert BL, Lebel RM, Lebel C. A multiparametric analysis of white matter maturation during late childhood and adolescence. Hum Brain Mapp. 2019;40(15):4345–4356.

40. Genc S, Malpas CB, Holland SK, Beare R, Silk TJ. Neurite density index is sensitive to age related differences in the developing brain. Neuroimage. 2017;148:373–380.

41. Dubois J, Dehaene-Lambertz G, Kulikova S, Poupon C, Huppi PS, Hertz-Pannier L. The early development of brain white matter: a review of imaging studies in fetuses, newborns and infants. Neuroscience. 2014;276:48–71.

42. Luders E, Thompson PM, Toga AW. The development of the corpus callosum in the healthy human brain. J Neurosci. 2010;30(33):10985–10990.

43. Wood NS, Costeloe K, Gibson AT, et al. The EPICure study: associations and antecedents of neurological and developmental disability at 30 months of age following extremely preterm birth. Arch Dis Child Fetal Neonatal Ed. 2005;90(2):F134–140.

44. Genc S, Seal ML, Dhollander T, Malpas CB, Hazell P, Silk TJ. White matter alterations at pubertal onset. Neuroimage. 2017;156:286–292.

